# Predicting Anxiety Trajectories from Individual Differences in Sleep–Related Distress Alleviation

**DOI:** 10.64898/2026.08.19.26360781

**Authors:** Lucija Blaževski, Sven Leach, Alejandro Osorio-Forero, Roy Cox, Joyce E. Reesen, Roxanne Bongers, Alex van Keeken, Savannah Ikelaar, Eus J.W. van Someren, Lara Rösler

**Affiliations:** Department of Sleep and Cognition, Netherlands Institute for Neuroscience (NIN), an Institute of the Royal Netherlands Academy of Arts and Sciences, Amsterdam, the Netherlands; Department of Sleep and Dreams, Netherlands Institute for Neuroscience (NIN), an Institute of the Royal Netherlands Academy of Arts and Sciences, Amsterdam, the Netherlands; Department of Integrative Neurophysiology, Center for Neurogenomics and Cognitive Research (CNCR), Amsterdam Neuroscience, Vrije Universiteit Amsterdam, the Netherlands; Department of Psychiatry, Amsterdam Public Health Research Institute and Amsterdam Neuroscience Research Institute, Amsterdam UMC, Vrije Universiteit, the Netherlands

## Abstract

**Importance:** Anxiety of fluctuating severity is common in many psychiatric disorders. Few studies addressed factors determining individual differences in trajectories of the recovery phase, while their identification could inspire treatment innovation. Given the role of REM sleep in overnight alleviation of emotional distress, we here investigate whether individual differences in this overnight regulatory process matter for anxiety recovery.

**Objective:** To investigate whether individual differences in overnight alleviation of distress by REM sleep predict anxiety recovery rate.

**Design:** People tend to volunteer for intervention trials when fluctuating symptoms peak. This results in a significant recovery even in waitlist or control conditions. Leveraging this opportunity to recruit people prior to the recovery phase, in this cohort study, we utilized data from people who volunteered for optional sleep EEG and overnight distress assessment prior to their participation in an intervention trial (2021-2025). Anxiety severity was assessed at baseline and two months later.

**Setting:** Home-based assessment in the Netherlands.

**Participants:** Adults with insomnia alongside cross-threshold symptom severities of generalized anxiety disorder, social anxiety disorder, panic disorder, posttraumatic stress disorder, or borderline personality disorder (N = 223, 157 female [70.4%]; mean [SD] age, 45.7 [14.5] years; clinical diagnoses confirmed in 165 [74.0%]).

**Exposures:** Cognitive behavioral therapy for insomnia (CBT-I) or waitlist control.

**Main Outcomes and Measures:** Predicting 2-month anxiety improvement by individual differences in the strength of the effect of REM sleep on overnight distress alleviation at baseline.

**Results:** Within-subject mixed model analysis showed stronger overnight distress alleviation across nights with longer REM sleep (*b* = -0.011; 95% CI, -0.016 to -0.007; *P* < .001). Individual differences in the strength of REM–related distress alleviation predicted anxiety improvement after two months (*b* = −0.521; 95% CI, −0.854 to −0.188; *P* = .002), irrespective of treatment or waitlist control (interaction *b* = 0.011; 95% CI, −0.656 to 0.678; *P* = .97).

**Conclusions and Relevance:** Individual differences in the degree to which REM sleep drives overnight alleviation of distress predict the trajectory of anxiety recovery in people with clinically relevant psychiatric complaints. These findings suggest REM-related emotion regulation as a mechanism linking sleep physiology to anxiety recovery.

**Key Points:** *Question:* Do individual differences in the degree to which sleep alleviates distress overnight predict the subsequent trajectory of anxiety severity?

*Findings:* In this cohort study, EEG recordings and measures of overnight distress alleviation were acquired across 662 nights in 223 adults experiencing anxiety related to different types of psychiatric conditions. Individual differences in the role of REM sleep in overnight distress alleviation predicted anxiety symptom improvement over 2 months.

*Meaning:* REM sleep–dependent overnight distress alleviation may represent a biomarker of anxiety recovery potential, highlighting REM-related emotional regulation as a candidate mechanism linking sleep physiology to clinical improvement.

## Main Text

Anxiety is common in many psychiatric conditions. Symptoms rarely follow a stable course but instead fluctuate over time. People tend to volunteer for intervention trials when fluctuating symptoms are at their peak and are likely to start improving intrinsically. Partly for this reason, clinical trials for anxiety frequently observe meaningful symptom reduction not only in active treatment arms but also among participants assigned to waitlist or control conditions.^1,2^ Yet the factors that determine why symptoms persist in some individuals but resolve in others, even in the absence of treatment, remain poorly understood.^3^ Uncovering the sources of this individual difference could help identify processes that shape the natural course of anxiety and, ultimately, inform treatment innovation. To date, efforts aimed at predicting the course of anxiety have largely focused on clinical features, while considerably less is known about physiological processes that may contribute to the resolution of anxiety over time.

Physiological processes taking place during sleep may be especially relevant. Poor sleep due to insomnia is highly comorbid with anxiety and is now recognized as a core feature rather than merely a secondary symptom.^4^ Importantly, insomnia severity contributes to functional impairment and poor psychiatric outcomes.^5,6^ Yet sleep is more than a correlate of psychopathology: unperturbed sleep has been proposed to actively support emotional regulation.^7,8^ Consistent with this role, nights of worse-than-usual sleep are followed by worse next-day affect within individuals.^9^

Rapid eye movement (REM) sleep seems particularly relevant for resolving emotional distress overnight. Processing of emotional experiences may benefit from the unique neurobiological conditions the brain experiences during REM sleep, such as prolonged periods of low noradrenaline levels.^10–12^ Experimental and neuroimaging studies confirmed a role of REM sleep in overnight adaptation of affective brain responses, including amygdala reactivity to previously encountered emotional material.^12,13^ The perturbed sleep that is characteristic of people with insomnia and/or anxiety disorders interferes with the benefits sleep could bring for emotion regulation.^12,13^ Most studies so far have focused on group averages of associations between sleep and emotional functioning, overlooking the possibility that there may be relevance in the between-subject variability in the contribution of REM sleep for the overnight resolution of distress.

Individual differences in the degree to which REM sleep supports overnight emotional regulation could be especially relevant for individual differences in anxiety symptom trajectories. An individual for whom a night with more REM sleep is consistently followed by a stronger overnight reduction in emotional distress has a greater capacity for sleep-dependent emotional regulation than an individual with little additional benefit from more REM sleep. Individual differences in REM sleep benefits likely extend beyond the overnight scale. Across nights, they could contribute to individual differences in recovery from anxiety.

We examined this possibility in adults with insomnia next to other clinically significant psychiatric conditions who volunteered for home-based sleep electroencephalography (EEG) and evening-to-morning distress assessments prior to entering a randomized trial on cognitive-behavioral therapy for insomnia (CBT-I). We first tested whether within-subject variation in REM sleep and in overnight distress alleviation were associated in this large sample of affected people. We then examined whether individual differences in REM-related distress alleviation predicted anxiety symptom improvement over the following 2 months. Given the participation of volunteers in a randomized trial design, ancillary analysis evaluated whether individual differences in REM sleep benefits matter for the degree of anxiety relief induced by insomnia treatment.

## Methods Study

### Design

Since people tend to volunteer for intervention trials when fluctuating anxiety symptoms peak and are likely to start improving again, we leveraged sleep EEG and overnight distress data that were optionally collected from a subset of participants in a randomized clinical trial (RCT). The RCT compares guided digital CBT-I with a waitlist control condition consisting of sleep diary monitoring only. The present study utilizes anxiety severity assessments collected at baseline (T0) and 2-month follow-up (T1) as part of that trial. Between T0 and T1, participants randomized to CBT-I completed the 5- to 8-week intervention; consequently, T1 in this group occurred 0 to 3 weeks after intervention completion. Data were collected from 2021 to 2025. The detailed trial protocol is described elsewhere.^14^ This study was approved by the Medical Ethical Committee of the VU University Medical Center (Registration number 2021/093) and registered in the International Clinical Trial Registry Platform (NL9776).

### Participants

A total of 291 adults volunteered for optional sleep EEG and overnight distress data collection. Participation in the RCT required them to have an Insomnia Severity Index (ISI)^15^ score ≥ 10, and clinically relevant symptoms of at least one of the following conditions that share anxiety as a key feature: generalized anxiety disorder (GAD), social anxiety disorder (SAD), panic disorder (PD), and posttraumatic stress disorder (PTSD), assessed using the Rapid Measurement Toolkit 20 (RMT20)^16^, or borderline personality disorder (BPD), assessed using the Ultrashort BPD Checklist (BPD-C).^17^ Clinical relevance was defined as a score at or above the cutoff on at least one subscale: GAD ≥ 11, SAD ≥ 12, PD ≥ 9, PTSD ≥ 8, or BPD ≥ 14. Diagnostic status for GAD, SAD, PD, and PTSD was additionally assessed using the Mini International Neuropsychiatric Interview^18^ (MINI). Exclusion criteria included alcohol or substance dependency, a current bipolar or psychotic disorder, and CBT-I within the past three months. Participants provided written or electronic informed consent and received financial compensation.

The within-person analysis of overnight distress alleviation required at least two eligible nights per participant, to allow reliable estimation of individual differences in the association between REM sleep and overnight distress change. Of the 291 participants (730 nights in total), 223 met this criterion and were included in the within-person analysis, contributing 662 nights. Of these 223 participants, 193 had GAD severity available at the 2-month follow-up and were included in the prognostic analysis of REM-related distress alleviation and subsequent anxiety severity. Sample characteristics are presented in Table 1, with baseline characteristics by treatment condition provided in eTable 1 in Supplement 1. GAD was the most prevalent diagnosis by far (see Table 1), consistent with its frequent co-occurrence with the other assessed conditions that share anxiety as a key feature ^19,20^. At least one clinical diagnosis of relevant psychiatric conditions was confirmed in 165 [74.0%] participants.

**Table 1.** Participant Characteristics (*N* = 223)

| **Characteristic** |  |
| --- | --- |
| **Demographics** |  |
| Age, mean (SD) | 45.7 (14.5) |
| Female, No. (%)<sup>a</sup> | 157 (70.4) |
| **Insomnia severity** |  |
| ISI total, mean (SD) | 17.2 (4.4) |
| **RMT20 subscale, mean (SD)** |  |
| Generalized anxiety disorder | 12.6 (3.3) |
| Social anxiety disorder | 10.9 (4.1) |
| Panic disorder | 7.2 (3.2) |
| Posttraumatic stress disorder | 8.8 (4.6) |
| **BPD-C severity, mean (SD)** |  |
| Borderline personality disorder | 19.1 (6.6) |
| **MINI-confirmed clinical diagnoses, No. (%)** |  |
| Generalized anxiety disorder | 132 (59.2) |
| Social anxiety disorder | 68 (30.5) |
| Panic disorder | 32 (14.3) |
| Posttraumatic stress disorder | 62 (27.8) |
| Any of the above diagnoses | 165 (74.0) |

Abbreviations: ISI, Insomnia Severity Index; MINI, Mini International Neuropsychiatric Interview; RMT20, Rapid Measurement Toolkit 20; BPD-C, Ultrashort BPD Checklist; SD, standard deviation.
<sup>a</sup> Sex was self-reported using the binary response options male or female; gender identity was not assessed separately.

### Procedure

#### Overnight distress and sleep recordings

At baseline, participants completed up to four consecutive nights of home-based sleep EEG recordings preceded and followed by assessment of emotional distress in a standardized context (Figure 1). Sleep EEG was recorded with the ambulatory EEG headband (ZMax Lite, Hypnodyne). This device measures two forehead EEG derivations (F7-Fpz, F8-Fpz), an accelerometer, and photoplethysmography, with a sampling frequency of 256 Hz. Before and after sleep, emotional distress was assessed with the validated Stress NRS-11 rating scale.^21^ To minimize contextual nuisance of the ratings, participants were instructed to provide each distress assessment in the same quiet location in their home, directly after completing a resting-state task (3 minutes eyes-closed followed by 3 minutes eyes-open). Instructions, timing, and assessments were done online through the Netherlands Sleep Registry (www.slaapregister.nl). This evening-night-morning sequence was repeated across consecutive nights, yielding paired within-person observations of pre-sleep distress, objective sleep architecture, and post-sleep distress.

**Figure 1.**
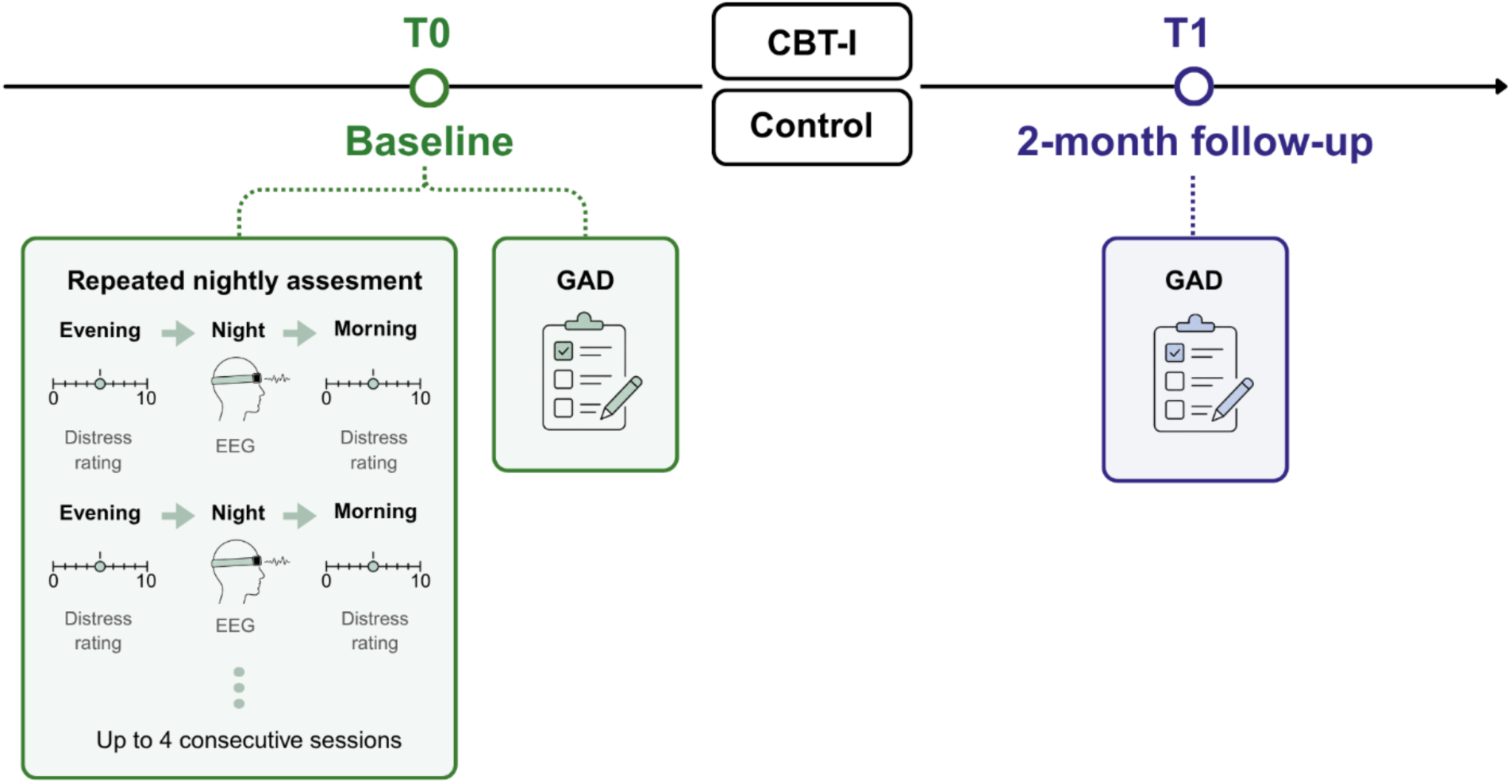
Study Design. Abbreviations: CBT-I, cognitive behavioral therapy for insomnia; EEG, electroencephalography; GAD, Generalized anxiety disorder measured using the Rapid Measurement Toolkit 20; T0, baseline; T1, 2-month follow-up. Following baseline, participants were randomized to guided digital CBT-I or a waitlist control condition.

#### Clinical assessment over 2 months

Generalized anxiety disorder symptom severity was assessed using the GAD subscale of the RMT20 at baseline and 2-month follow-up.^16^ The SAD, PD, and PTSD subscales of the RMT20, together with BPD symptoms assessed using the BPD-C,^17^ were included as secondary psychiatric symptom dimensions.

### EEG Data Processing

Full details of signal quality check, sleep staging, and sleep opportunity detection are provided in the eMethods in Supplement 1. All sleep data processing was performed in MATLAB (version 2022b).^22^ Baseline sleep architecture measures are presented in eTable 2 in Supplement 1. Sleep staging was performed semi-automatically in 30-second epochs, combining two validated deep learning models: uSleep^23^ and SleepTransformer,^24^ using soft vote weighting^25^. Validation of this approach against manual sleep staging is shown in eFigure 1 in Supplement 1. All resulting scores were checked by an expert scorer. REM sleep duration was derived from sleep scores using SleepTrip.^26^

### Analysis

Statistical analyses were conducted in R (version 4.4.2)^27^ using the lme4 (version 1.1.37)^28^ and lmerTest (version 3.1.3)^29^ packages for multilevel modeling. All statistical tests were 2-sided, with a significance threshold of α = .05. Analyses were based on complete cases for the variables included in each model; no missing data were imputed. To isolate within-person effects, time-varying predictors, including REM duration and evening distress, were centered around each participant’s mean across sessions.

To test whether REM sleep predicted overnight distress alleviation and to derive an individualized index of this capacity, we estimated a linear mixed-effects model among participants with at least two eligible nights. Next-morning distress was modeled as a function of within-person-centered REM duration, adjusting for evening distress, with a random intercept and a random slope for REM duration by participant. The fixed effect of REM duration tested the average within-person association, while the participant-specific random slopes provided an individualized estimate of each participant’s REM–distress association. More negative slopes indicated that nights with longer REM sleep than usual were followed by greater overnight alleviations in distress. Participant-specific REM slopes were standardized and reverse-coded so that higher values represented stronger REM-linked overnight distress alleviation.

We then tested whether individual differences in REM-related overnight distress alleviation predicted GAD severity at the 2-month follow-up, beyond baseline GAD severity. The model additionally included treatment condition and a REM-slope-by-treatment interaction to determine whether the prognostic value of the REM slope differed between CBT-I and the control condition. To explore whether individual differences in REM-related overnight distress alleviation were associated with follow-up symptom severity beyond GAD, we repeated the analysis using SAD, PD, PTSD, and BPD symptom severity as secondary outcomes.

## Results

### REM Sleep and Overnight Distress Alleviation

More REM sleep on a given night was associated with greater overnight alleviation of distress (*b* = -0.011; 95% CI, -0.016 to -0.007; *P* < .001; Figure 2A). The association with REM sleep was the strongest observed across the sleep architecture and continuity measures examined (eTable 3 in Supplement 1).

**Figure 2.**
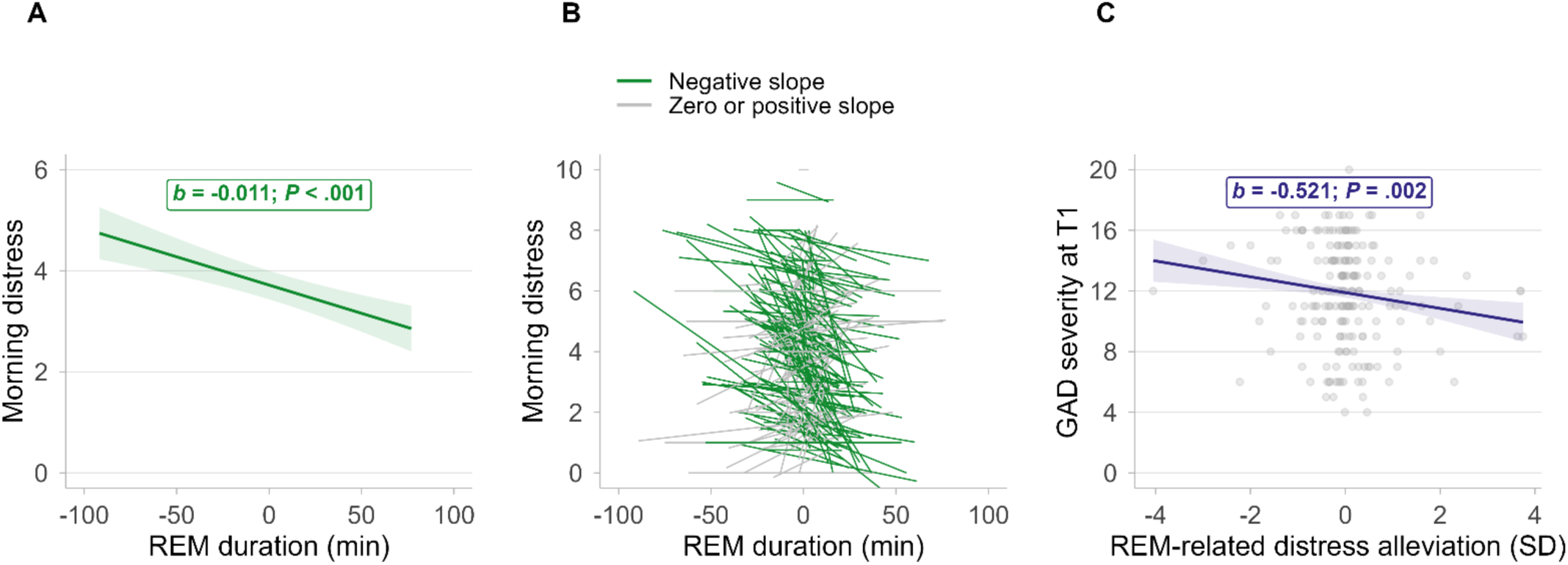
REM Sleep and Distress Alleviation From Nightly Variation to Follow-up Anxiety. Abbreviations: GAD, generalized anxiety disorder; REM, rapid eye movement sleep; SD, standard deviation; T1, 2-month follow-up. A, Association between within-person-centered REM duration in minutes (x-axis) and next-morning distress (y-axis), estimated from a linear mixed-effects model controlling for evening distress. The shaded region indicates the 95% confidence interval. B, Individual variability in REM-related overnight distress alleviation. Each line represents one participant’s unadjusted linear fit of next-morning distress on REM duration, reflecting the change in next-morning distress per additional minute of REM sleep. Green lines indicate negative slopes, reflecting greater overnight distress alleviation with longer REM duration, whereas gray lines indicate flat or positive slopes, reflecting unchanged or increased morning distress with longer REM duration. C, Association between individual REM-related overnight distress alleviation at baseline and GAD severity at 2-month follow-up (*n* = 193). Participant-specific REM slopes were standardized (x-axis in SD units) and reverse-coded so that higher values indicated stronger REM-linked overnight distress alleviation; each point represents one participant. The shaded region indicates the 95% confidence interval.

### REM-Related Distress Alleviation Predicts Follow-up Anxiety Severity

The extent to which nights with more REM sleep were followed by overnight resolution of distress varied across individuals (Figure 2B). Greater individual REM-related overnight distress alleviation at baseline was associated with lower GAD severity 2 months later, after adjustment for baseline GAD severity (*b* = −0.521; 95% CI, −0.854 to −0.188; *P* = .002; Figure 2C). Thus, individuals who showed greater distress alleviation on nights with more REM sleep also showed lower subsequent anxiety severity. This association was similar across CBT-I and control conditions (interaction *b* = 0.011; 95% CI, −0.656 to 0.678; *P* = .97). Complete model estimates are provided in eTable 4 in Supplement 1. In secondary analyses, greater REM-related overnight distress alleviation was also associated with lower SAD and PD at follow-up, whereas associations with PTSD and BPD symptoms were not significant (eTable 5 in Supplement 1).

## Discussion

In this study of individuals with clinically significant insomnia and co-occuring psychiatric conditions, nights with longer REM sleep were followed by stronger overnight alleviation of distress. Importantly, the strength of this relationship varied between individuals, and greater REM-related overnight distress alleviation at baseline was associated with lower anxiety severity two months later. Together, these findings suggest that individual differences in sleep-dependent emotional regulation carry prognostic information about subsequent anxiety course.

Our findings align with experimental and theoretical work linking REM sleep to emotional regulation.^11–13,30^ Previous studies have largely examined these processes under controlled experimental conditions; here, night-to-night variation in REM sleep was associated with alleviation of distress in the daily lives of individuals with insomnia and co-occurring psychiatric conditions. Crucially, the clinically relevant finding was the marked inter-individual variability in the REM–distress association, beyond the average relationship. Between-person differences in overnight distress alleviation may become consequential over longer timescales: repeated failure to obtain overnight relief could allow emotional distress to carry forward across days; more effective overnight alleviation may create conditions for symptoms to diminish over time. Repeated variation in this nightly regulatory process may therefore accumulate into meaningful differences in longer-term anxiety course, consistent with our finding that stronger REM-related distress alleviation predicted lower subsequent anxiety.

The prospective association with anxiety severity is particularly relevant given the marked heterogeneity in the trajectory of anxiety symptoms. Hovenkamp-Hermelink and colleagues^3^ showed that anxiety disorders can follow widely divergent courses. They identified several clinical and psychological characteristics associated with persistence, but the contribution of biological characteristics remained considerably less clear. More recent work similarly indicates that reliable individual-level prediction of longitudinal anxiety trajectories remains underdeveloped.^31^ Our findings point to a complementary source of prognostic information: REM-related overnight distress alleviation which captures a physiological process repeatedly expressed within an individual. Because the strength of REM-related overnight distress alleviation can be derived from wearable EEG and distress assessment at home, the measure offers a scalable means of characterizing overnight emotional-regulatory capacity. This perspective aligns with Goldstein-Piekarski and colleagues,^32^ who highlighted the potential value of integrating sleep physiology and wearable technologies into more individualized approaches to mood and anxiety disorders. Thus, variation in sleep-dependent emotional regulation may represent a biomarker of heterogeneity in anxiety course that is not captured by conventional clinical characteristics alone.

That the prognostic association did not differ by treatment condition further suggests that REM-related overnight distress alleviation may characterize anxiety course more broadly, rather than response to CBT-I specifically. Nevertheless, this may help explain why improvements in insomnia symptoms following CBT-I do not translate uniformly into anxiety relief.^33^ CBT-I effects may primarily operate through behavioral and cognitive pathways, such as reducing conditioned arousal and correcting maladaptive sleep behaviors, without necessarily modifying the neurobiological mechanisms through which REM sleep contributes to overnight emotional recovery. Consistent with a potential dissociation between insomnia improvement and REM physiology, a recent multicenter investigation found that CBT-I improved insomnia-related outcomes without altering REM duration.^34^ The same framework may explain why robust pretreatment moderators of anxiety response to CBT-I have been difficult to identify.^33^

What might determine the capacity of REM sleep to support overnight distress alleviation? One candidate mechanism is variability in noradrenergic suppression during this stage. Under healthy conditions, locus coeruleus activity is strongly reduced during REM sleep,^10^ creating conditions thought to support emotional processing. If this reduction fails, for example, in the context of chronic distress or hyperarousal, REM sleep may be less effective in supporting overnight emotional recalibration.^30^ Further research that manipulates noradrenergic activity during sleep could clarify whether differences in REM-related noradrenergic silencing explain why sleep supports overnight distress alleviation in some individuals more than in others.

The present findings should be interpreted in light of several limitations. First, distress was operationalized using a single subjective rating to improve adherence across repeated multi-day assessments. Although this rating captured within-person fluctuations, the measure does not distinguish between cognitive, affective, or physiological components of distress. Future studies should test whether our findings extend to these different components. Additionally, our sample included individuals with insomnia and other co-occurring psychiatric conditions (including GAD, PTSD, BPD, PD, and SAD) and both clinical and subclinical severity, which strengthens confidence that our findings are not specific to a single diagnostic group or symptom severity. Nonetheless, whether REM-related overnight distress alleviation generalizes to individuals with psychiatric disorders without insomnia remains to be explored.

Overall, we provide evidence that REM-related overnight distress alleviation varies meaningfully between individuals with insomnia and co-occurring emotional disorders and is prospectively associated with subsequent anxiety severity. By linking a repeatedly expressed nightly process to subsequent symptom course, our results identify sleep-dependent emotional regulation as a candidate physiological contributor to heterogeneity in anxiety trajectories. More broadly, we demonstrate that understanding anxiety trajectories may require considering not only the severity of symptoms while awake, but also the processes through which emotional distress is - or is not - resolved during sleep.

## Data Availability

All data produced in the present study are available upon reasonable request to the authors

## Acknowledgments

**Data Access, Responsibility, and Analysis:** Dr Rösler and Ms Blaževski had full access to all the data in the study and take responsibility for the integrity of the data and the accuracy of the data analysis.

**Data Sharing Statement:** The data supporting the findings of this study are available from the corresponding author upon reasonable request. Analysis code is available at https://github.com/lucija-blazevski/Sleep-Stress-Coupling.

## Author contributions

**Concept and design:** Blaževski, van Someren, Rösler.

**Data acquisition:** Reesen, Bongers, van Keeken, Ikelaar.

**Analysis or interpretation of data:** Blaževski, Leach, Osorio-Forero, Cox, Rösler.

**Drafting of the manuscript:** Blaževski.

**Critical review of the manuscript for important intellectual content**: All authors.

**Statistical analysis:** Blaževski, Rösler.

**Obtained funding:** van Someren, Rösler.

**Administrative, technical, or material support:** Osorio-Forero, Cox, Reesen, Bongers, van Keeken, Ikelaar.

**Supervision:** Rösler.

**Conflict of Interest Disclosures:** Dr Cox is an employee of Deep Sleep Technologies; his contribution to this study was completed before the start of this employment, and Deep Sleep Technologies had no role in the study. The other authors have no conflicts of interest to declare.

**Funding/Support:** This study was supported by BIAL (253/2022) grant awarded to Dr Rösler and Hersenstichting (DR-2019-00345 and DR-2019-00322), ZonMw Leefstijlgeneeskunde (555003203), and European Research Council Advanced (101055383 OVERNIGHT) grants awarded to Dr van Someren. Views and opinions expressed are however those of the author(s) only and do not necessarily reflect those of the European Union or the European Research Council Executive Agency. Neither the European Union nor the granting authority can be held responsible for them.

**Role of the Funder/Sponsor:** The funders had no role in the design and conduct of the study; collection, management, analysis, and interpretation of the data; preparation, review, or approval of the manuscript; and decision to submit the manuscript for publication.

**Use of Artificial Intelligence:** OpenAI ChatGPT and Anthropic Claude were used to assist with text and code editing. The authors reviewed all AI-assisted content and take full responsibility for the accuracy of the final manuscript.

## References

1. Motta LS, Gosmann NP, Costa M de A, et al. Placebo response in trials with patients with anxiety, obsessive-compulsive and stress disorders across the lifespan: a three-level meta-analysis. BMJ Ment Health. 2023;26(1):e300630. doi:10.1136/bmjment-2022-300630

2. Scott AJ, Bisby MA, Heriseanu AI, et al. Understanding the untreated course of anxiety disorders in treatment-seeking samples: A systematic review and meta-analysis. J Anxiety Disord. 2022;89:102590. doi:10.1016/j.janxdis.2022.102590

3. Hovenkamp-Hermelink JHM, Jeronimus BF, Myroniuk S, Riese H, Schoevers RA. Predictors of persistence of anxiety disorders across the lifespan: a systematic review. Lancet Psychiatry. 2021;8(5):428–443. doi:10.1016/S2215-0366(20)30433-8

4. Cox RC, Olatunji BO. A systematic review of sleep disturbance in anxiety and related disorders. J Anxiety Disord. 2016;37:104–129. doi:10.1016/j.janxdis.2015.12.001

5. Neckelmann D, Mykletun A, Dahl AA. Chronic Insomnia as a Risk Factor for Developing Anxiety and Depression. Sleep. 2007;30(7):873–880. doi:10.1093/sleep/30.7.873

6. Soehner AM, Harvey AG. Prevalence and Functional Consequences of Severe Insomnia Symptoms in Mood and Anxiety Disorders: Results from a Nationally Representative Sample. Sleep. 2012;35(10):1367–1375. doi:10.5665/sleep.2116

7. Goldstein AN, Walker MP. The Role of Sleep in Emotional Brain Function. Annu Rev Clin Psychol. 2014;10:679–708. doi:10.1146/annurev-clinpsy-032813-153716

8. Tempesta D, Socci V, De Gennaro L, Ferrara M. Sleep and emotional processing. Sleep Med Rev. 2018;40:183–195. doi:10.1016/j.smrv.2017.12.005

9. Bourke M, Harrison Z, Staton S, Rossa K, Smith S. Sleep well, feel well and vice versa? A meta-analysis of daily bidirectional within-person associations between sleep and affect. Sleep Med Rev. 2026;86:102232. doi:10.1016/j.smrv.2026.102232

10. Aston-Jones G, Bloom FE. Activity of norepinephrine-containing locus coeruleus neurons in behaving rats anticipates fluctuations in the sleep-waking cycle. J Neurosci. 1981;1(8):876–886. doi:10.1523/JNEUROSCI.01-08-00876.1981

11. Poe GR. Sleep Is for Forgetting. J Neurosci. 2017;37(3):464–473. doi:10.1523/JNEUROSCI.0820-16.2017

12. van der Helm E, Yao J, Dutt S, Rao V, Saletin JM, Walker MP. REM Sleep Depotentiates Amygdala Activity to Previous Emotional Experiences. Curr Biol. 2011;21(23):2029–2032. doi:10.1016/j.cub.2011.10.052

13. Wassing R, Lakbila-Kamal O, Ramautar JR, Stoffers D, Schalkwijk F, Someren EJWV. Restless REM Sleep Impedes Overnight Amygdala Adaptation. Curr Biol. 2019;29(14):2351–2358.e4. doi:10.1016/j.cub.2019.06.034

14. Reesen JE, Van Der Zweerde T, Batelaan NM, et al. Do better nights lead to better days? Guided internet-based cognitive behavioral therapy for insomnia in people suffering from a range of mental health problems: Protocol of a pragmatic randomized clinical trial. Contemp Clin Trials. 2023;127:107122. doi:10.1016/j.cct.2023.107122

15. Morin CM, Belleville G, Bélanger L, Ivers H. The Insomnia Severity Index: psychometric indicators to detect insomnia cases and evaluate treatment response. Sleep. 2011;34(5):601–608. doi:10.1093/sleep/34.5.601

16. Batterham PJ, Sunderland M, Carragher N, Calear AL. Development of the RMT20, a composite screener to identify common mental disorders. BJPsych Open. 2020;6(3):e50. doi:10.1192/bjo.2020.37

17. Bloo J, Arntz A, Schouten E. The Borderline Personality Disorder Checklist: Psychometric evaluation and factorial structure in clinical and nonclinical samples. Ann Psychol. 2017;20(2):311–336. doi:10.18290/rpsych.2017.20.2-3en

18. Sheehan DV, Lecrubier Y, Sheehan KH, et al. The Mini-International Neuropsychiatric Interview (M.I.N.I.): the development and validation of a structured diagnostic psychiatric interview for DSM-IV and ICD-10. J Clin Psychiatry. 1998;59 Suppl 20:22-33;quiz 34–57.

19. Brown TA, Campbell LA, Lehman CL, Grisham JR, Mancill RB. Current and lifetime comorbidity of the DSM-IV anxiety and mood disorders in a large clinical sample. J Abnorm Psychol. 2001;110(4):585–599. doi:10.1037//0021-843x.110.4.585

20. Goldstein-Piekarski AN, Williams LM, Humphreys K. A trans-diagnostic review of anxiety disorder comorbidity and the impact of multiple exclusion criteria on studying clinical outcomes in anxiety disorders. Transl Psychiatry. 2016;6(6):e847–e847. doi:10.1038/tp.2016.108

21. Karvounides D, M. Simpson P, Davies WH, A. Khan K, J. Weisman S, R. Hainsworth K. Three studies supporting the initial validation of the stress numerical rating scale-11 (Stress NRS-11): A single item measure of momentary stress for adolescents and adults. Pediatr Dimens. 2016;1(4). doi:10.15761/PD.1000124

22. MATLAB. Published online 2022. https://www.mathworks.com

23. Perslev M, Darkner S, Kempfner L, Nikolic M, Jennum PJ, Igel C. U-Sleep: resilient high-frequency sleep staging. Npj Digit Med. 2021;4(1):72. doi:10.1038/s41746-021-00440-5

24. Phan H, Mikkelsen K, Chen OY, Koch P, Mertins A, De Vos M. SleepTransformer: Automatic Sleep Staging With Interpretability and Uncertainty Quantification. IEEE Trans Biomed Eng. 2022;69(8):2456–2467. doi:10.1109/TBME.2022.3147187

25. Salfi F, Corigliano D, Amicucci G, et al. The potential of ensemble-based automated sleep staging on single-channel EEG signal from a wearable device. bioRxiv. Preprint posted online July 24, 2025:2025.07.21.665164. doi:10.1101/2025.07.21.665164

26. Cox R, Weber FD, Van Someren EJW. Customizable automated cleaning of multichannel sleep EEG in SleepTrip. Front Neuroinformatics. 2024;18:1415512. doi:10.3389/fninf.2024.1415512

27. R Core Team. R: A Language and Environment for Statistical Computing. Published online 2024. Accessed April 27, 2026. https://www.r-project.org/

28. Bates D, Mächler M, Bolker B, Walker S. Fitting Linear Mixed-Effects Models Using lme4. J Stat Softw. 2015;67:1–48. doi:10.18637/jss.v067.i01

29. Kuznetsova A, Brockhoff PB, Christensen RHB. lmerTest Package: Tests in Linear Mixed Effects Models. J Stat Softw. 2017;82:1–26. doi:10.18637/jss.v082.i13

30. Van Someren EJW. Brain mechanisms of insomnia: new perspectives on causes and consequences. Physiol Rev. 2021;101(3):995–1046. doi:10.1152/physrev.00046.2019

31. Fairweather SJ, Fraser H, Lam N, et al. Prediction models for longitudinal trajectories of depression and anxiety: a systematic review. J Affect Disord. 2026;401:121255. doi:10.1016/j.jad.2026.121255

32. Goldstein-Piekarski AN, Holt-Gosselin B, O’Hora K, Williams LM. Integrating sleep, neuroimaging, and computational approaches for precision psychiatry. Neuropsychopharmacology. 2020;45(1):192–204. doi:10.1038/s41386-019-0483-8

33. Mirchandaney R, Barete R, Asarnow LD. Moderators of Cognitive Behavioral Treatment for Insomnia on Depression and Anxiety Outcomes. Curr Psychiatry Rep. 2022;24(2):121–128. doi:10.1007/s11920-022-01326-3

34. Sforza M, Morin CM, Dang-Vu TT, et al. The effectiveness of Cognitive behavioral therapy for insomnia on sleep EEG hyperarousal: a multicentric polysomnographic study. Transl Psychiatry. 2026;16(1):88. doi:10.1038/s41398-026-03882-1

