## Supplemental Material for "Predicting Anxiety Trajectories from Individual Differences in Sleep–Related Distress Alleviation"

**Supplement 1.**

eTable 1. Baseline Characteristics by Treatment Condition

eMethods. EEG Signal Quality Assessment, Sleep Staging, and Sleep Opportunity Detection

eFigure 1. Confusion Matrix for Automated Sleep Staging Against Manual Expert Scoring

eResults. Additional Sleep and Clinical Outcome Analyses

eTable 2. Baseline Sleep Architecture and Sleep-Continuity Measures by Treatment Condition

eTable 3. Within-Person Associations Between Sleep Variables and Overnight Distress Alleviation

eTable 4. Model Estimates for GAD Severity at 2-Month Follow-up

eTable 5. Association Between Baseline REM-Related Overnight Distress Alleviation and Symptom Severity at 2-Month Follow-up

eReferences

### eTable 1. Baseline Characteristics by Treatment Condition

| Characteristic | CBT-I *n* = 111 | Control *n* = 112 |
|---|---|---|
| **Demographics** | | |
| Age, mean (SD) | 43.8 (14.3) | 47.6 (14.5) |
| Female, No. (%)[a] | 76 (68.5) | 81 (72.3) |
| **Insomnia severity** | | |
| ISI total, mean (SD) | 17.2 (4.2) | 17.2 (4.5) |
| **RMT20 subscale, mean (SD)** | | |
| Generalized anxiety disorder | 12.8 (3.4) | 12.4 (3.3) |
| Social anxiety disorder | 11.0 (4.0) | 10.8 (4.2) |
| Panic disorder | 7.2 (3.0) | 7.1 (3.4) |
| Posttraumatic stress disorder | 9.0 (4.6) | 8.7 (4.7) |
| **BPD-C severity, mean (SD)** | | |
| Borderline personality disorder | 19.9 (7.1) | 18.2 (5.8) |
| **MINI-confirmed clinical diagnoses, No. (%)** | | |
| Generalized anxiety disorder | 70 (63.1%) | 62 (55.4%) |
| Social anxiety disorder | 36 (32.4%) | 32 (28.6%) |
| Panic disorder | 14 (12.6%) | 18 (16.1%) |
| Posttraumatic stress disorder | 35 (31.5%) | 27 (24.1%) |
| Any of the above diagnoses | 85 (76.6%) | 80 (71.4%) |

Abbreviations: CBT-I, cognitive behavioral therapy for insomnia; ISI, Insomnia Severity Index; MINI, Mini International Neuropsychiatric Interview; RMT20, Rapid Measurement Toolkit 20; BPD-C, Ultrashort BPD Checklist; SD, standard deviation.

[a] Sex was self-reported using the binary response options male or female; gender identity was not assessed separately.

**EEG Signal Quality Assessment**

Before sleep staging, EEG signal quality was screened independently for each channel using two spectral criteria applied at the 10-second epoch level. First, excessive power was identified by calculating, for each 10-second epoch, the percentage of frequencies between 1 and 45 Hz for which a power spectral density was above 1000 $\mu V^2/Hz$, approximating manual identification of unusable data segments through time-frequency representations. This percentage was smoothed over time using a 170-second moving median, and epochs were flagged when the smoothed value exceeded 5%. Second, signal dropout was identified by calculating the percentage of frequencies between 1 and 20 Hz with a power spectral density below 0.1 $\mu V^2/Hz$. This measure was smoothed using a 50-second moving median, and epochs were flagged when the smoothed value exceeded 70%. Window lengths for excessive power and signal dropout were selected during piloting to best flag low-quality epochs without discarding clean segments. A 10-second epoch was considered artefactual for a given channel if it met either criterion. Artefact labels were then aggregated to the 30-second epoch level: a 30-second epoch was marked artefactual for a channel if any of its three constituent 10-second epochs was flagged as artefactual. Finally, a 30-second epoch was considered unscorable and excluded from sleep architecture calculations if it was marked artefactual in both channels simultaneously.

**Sleep Staging**

Sleep staging was performed semi-automatically in 30-second epochs. We used Sleepyland[1], an automated pipeline, and combined two validated deep learning models: uSleep[2] and SleepTransformer[3]. The models received both EEG channels as input and were applied in parallel to the raw and bandpass-filtered signal (0.5–35 Hz; zero-phase, two-pass Butterworth IIR filters, each $6^{th}$-order), comparable to a conventional polysomnography montage. Each model generated epoch-level hypnodensities, i.e., probability distributions across sleep stages, which were combined across the two EEG channels as their mean within Sleepyland. These channel-averaged hypnodensities from uSleep and SleepTransformer, each obtained for the raw and filtered signals, were combined using confidence-weighted soft voting.[4] Specifically, the four hypnodensities for each epoch were combined using a weighted mean, with the maximum stage probability within each hypnodensity used as its confidence weight. Consequently, outputs that more strongly favored a particular stage contributed more to the final consensus probabilities. To validate this automatic approach, we compared it against fully manual expert scoring on a subset of 237 nights, obtaining Cohen's kappa ($\kappa$) of 0.76 - in line with inter-rater reliability typically reported for sleep staging in the literature.[5] Stage-by-stage agreement was as follows: wake $\kappa = 0.83$, REM $\kappa = 0.82$, N3 $\kappa = 0.75$, N2 $\kappa = 0.72$, and N1 $\kappa = 0.24$ (see eFigure 1 for full confusion matrix). For these 237 nights, the fully manual scores were used in the final data set. For all remaining nights, the automatic consensus scores were reviewed by a trained scorer and corrected where necessary.

**Sleep Opportunity Detection**

The sleep opportunity window for each recording was determined using a semi-automated approach that combined accelerometer-based posture detection and resting-state timestamps, followed by a manual confirmation. We used the headband's built-in triaxial accelerometer to classify posture. Upright postures were detected using a threshold based on the 10th and 90th percentiles of the accelerometer's x-axis signal per recording. Sleep opportunity start was defined as the onset of the first lying bout of at least 5 minutes that began after the evening resting state period and before the first sleep epoch. Sleep opportunity end was defined as the onset of the first non-lying bout of at least 2 minutes, occurring after the last scored sleep epoch. If accelerometer-based detection failed due to noise or unclear transitions, timestamps from the evening and morning resting-state tasks were used. All automatically detected windows were reviewed and corrected as needed by a trained researcher.

### eFigure 1. Confusion Matrix for Automated Sleep Staging Against Manual Expert Scoring

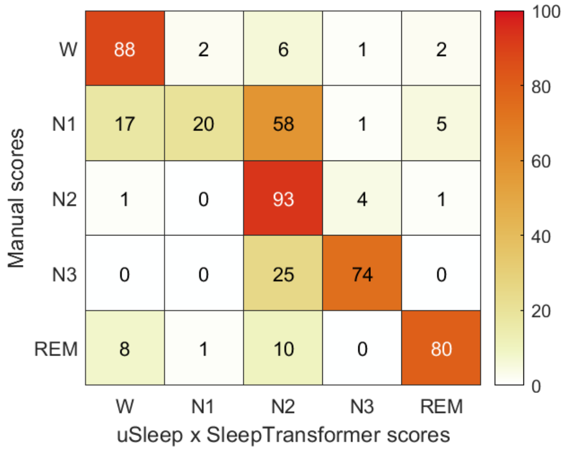


Abbreviations: N1, N2, and N3, non–rapid eye movement sleep stages 1, 2, and 3; REM, rapid eye movement sleep; W, wake. κ, Cohen κ coefficient. Values represent the mean percentage of epochs from each manually scored stage (rows) assigned to each automatically scored stage (columns), averaged across participants after within-participant normalization in 237 nights. Stage-by-stage agreement: wake κ = 0.83, REM κ = 0.82, N3 κ = 0.75, N2 κ = 0.72, N1 κ = 0.24.

### eResults

#### Baseline Sleep Characteristics

Baseline sleep architecture and sleep-continuity measures were averaged across each participant's available nights before calculating group-level descriptive statistics.

### eTable 2. Baseline Sleep Architecture and Sleep-Continuity Measures by Treatment Condition

| **Sleep variable** | **Total** (*N* = 223) | **CBT-I** (*n* = 111) | **Control** (*n* = 112) |
|---|---|---|---|
| N1 duration, min | 11.8 (8.6) | 11.9 (8.8) | 11.8 (8.5) |
| N2 duration, min | 212.3 (49.8) | 211.4 (52.7) | 213.2 (47.1) |
| N3 duration, min | 79.8 (30.6) | 81.0 (32.3) | 78.5 (28.9) |
| REM duration, min | 89.0 (23.5) | 88.5 (24.9) | 89.5 (22.1) |
| Total sleep time, min | 392.9 (60.7) | 392.8 (68.5) | 393.0 (52.1) |
| Sleep efficiency, % | 82.1 (9.7) | 82.1 (10.5) | 82.0 (8.8) |
| Wake after sleep onset, min | 50.0 (36.1) | 47.8 (33.6) | 52.3 (38.4) |
| Sleep onset latency, min | 20.8 (19.0) | 21.9 (19.8) | 19.7 (18.2) |

Abbreviations: CBT-I, cognitive behavioral therapy for insomnia; SD, standard deviation; N1, N2, and N3, non–rapid eye movement sleep stages 1, 2, and 3; REM, rapid eye movement sleep. Data are presented as the mean (SD) of participant-level averages across available nights.

**Comparison of Sleep Variables as Predictors of Overnight Distress Alleviation**

To assess whether the association with overnight distress alleviation was specific to REM sleep, we conducted exploratory analyses of other sleep architecture and sleep-continuity measures, including N1, N2, and N3 duration, total sleep time, sleep efficiency, wake after sleep onset, and sleep onset latency. Analyses were restricted to participants with at least 2 nights of usable data. For comparability across sleep measures, predictors were standardized within participants. For each sleep measure, morning distress was modeled as a function of the within-person sleep predictor and evening distress. Models included participant-specific random intercepts and random slopes for the sleep predictor. For sleep measures for which the random-slope specification did not yield convergence, a random-intercept-only model was used. REM was the prespecified primary sleep variable; the Holm-Bonferroni correction was applied across the 7 remaining variables.

REM duration showed the largest within-person association with overnight distress alleviation ($b = -0.273$; 95% CI, −0.401 to −0.146; $P < .001$). After Holm-Bonferroni correction, TST, SE, and N3 duration were also associated with overnight alleviation of distress, whereas N1 and N2 durations, WASO, and SOL were not (eTable 3). Because TST and SE partly comprise REM sleep, their associations cannot be interpreted as independent of REM; the weaker association of N3 duration indicates that the effect, while strongest for REM, was not exclusive to it.

### eTable 3. Within-Person Associations Between Sleep Variables and Overnight Distress Alleviation

| Sleep variable | *b* | 95% CI | *P* Value | $P_{\text{Holm}}$ Value |
|---|---|---|---|---|
| REM duration | −0.273 | [−0.401, −0.146] | < .001 | — |
| N1 duration | −0.054 | [−0.186, 0.078] | .42 | .85 |
| N2 duration | −0.142 | [−0.272, −0.013] | .03 | .13 |
| N3 duration | −0.173 | [−0.298, −0.048] | .007 | .04 |
| Total sleep time | −0.241 | [−0.365, −0.117] | < .001 | .001 |
| Sleep efficiency | −0.235 | [−0.362, −0.107] | < .001 | .002 |
| Wake after sleep onset | 0.135 | [−0.003, 0.273] | .06 | .17 |
| Sleep onset latency | 0.017 | [−0.110, 0.143] | .80 | .85 |

Abbreviations: *b*, regression coefficient per 1-SD within-person increase in the sleep variable; CI, confidence interval; REM, rapid eye movement. Sleep predictors were standardized within participants. Models included evening distress and participant-specific random intercepts and random slopes for the sleep predictor; for N3 duration, total sleep time, and sleep onset latency, the random-slope specification yielded singular fits, and random-intercept-only models were therefore used. Holm-Bonferroni correction was applied across the 7 secondary sleep measures; REM duration was not included.

### eTable 4. Model Estimates for GAD Severity at 2-Month Follow-up

| Variable | *b* | 95% CI | *P* Value |
|---|---|---|---|
| REM-related overnight distress alleviation, per 1 SD | −0.521 | [−0.854, −0.188] | .002 |
| Treatment condition (CBT-I vs control) | −0.488 | [−1.156, 0.179] | .15 |
| Baseline GAD severity | 0.749 | [0.650, 0.847] | < .001 |
| REM-related distress alleviation × treatment condition | 0.011 | [−0.656, 0.678] | .97 |

Abbreviations: *b* = coefficient; CBT-I = cognitive behavioral therapy for insomnia; CI = confidence interval; GAD = generalized anxiety disorder; REM = rapid eye movement; SD = standard deviation. Data include 193 participants. REM-related overnight distress alleviation was derived from participant-specific REM slopes, standardized and reverse-coded so that higher values indicate greater overnight distress alleviation on nights with more REM sleep. Treatment condition was effect-coded (control = −0.5; CBT-I = 0.5); therefore, the coefficient for REM-related distress alleviation represents its average association with follow-up GAD severity across treatment conditions.

**Secondary Analyses Across Anxiety Symptom Dimensions**

To assess whether the association between baseline REM-related overnight distress alleviation and clinical outcome extended beyond GAD, the analysis was repeated for social anxiety disorder (SAD), panic disorder (PD), posttraumatic stress disorder (PTSD), and borderline personality disorder (BPD) symptoms, with Holm-Bonferroni correction across the 4 secondary outcomes. Stronger REM-related overnight distress alleviation was associated with lower SAD and PD severity at the 2-month follow-up, whereas associations with PTSD and BPD symptoms were not significant (eTable 5). REM-related distress alleviation by treatment interactions were not significant for SAD (*b* = −0.289; 95% CI, −1.034 to 0.455), PD (*b* = −0.206; 95% CI, −0.821 to 0.410), PTSD (*b* = −0.420; 95% CI, −1.228 to 0.388), or BPD (*b* = 0.257; 95% CI, −0.811 to 1.325; all Holm-Bonferroni-adjusted *P* > .99).

### eTable 5. Association Between Baseline REM-related Overnight Distress Alleviation and Secondary Symptom Severity at 2-Month Follow-up

| Symptom Dimension | *b* | 95% CI | *P* Value | $P_{Holm}$ Value |
|---|---|---|---|---|
| Social anxiety disorder | -0.525 | [-0.894, -0.157] | .005 | .02 |
| Panic disorder | -0.485 | [-0.793, -0.178] | .002 | .009 |
| Posttraumatic stress disorder | -0.095 | [-0.498, 0.309] | .64 | .65 |
| Borderline personality disorder | -0.268 | [-0.803, 0.266] | .32 | .65 |

Abbreviations: *b,* regression coefficient; CI = confidence interval. Each coefficient was obtained from a separate linear regression model including baseline REM-related overnight distress alleviation, treatment condition, their interaction, and baseline severity of the corresponding symptom dimension. The reported coefficient represents the average association across treatment conditions. Sample sizes were 193 for social anxiety, panic disorder, and posttraumatic stress disorder, and 194 for borderline personality disorder. Participant-specific REM slopes were standardized and reverse-coded so that higher values indicated stronger REM-linked overnight distress alleviation; therefore, negative coefficients indicate that stronger REM-related overnight distress alleviation was associated with lower symptom severity at follow-up. Holm-Bonferroni correction was applied across the 4 secondary outcomes.

**eReferences**

1. Rossi AD, Metaldi M, Bechny M, et al. SLEEPYLAND: trust begins with fair evaluation of automatic sleep staging models. *Npj Digit Med.* 2025;9(1):55. doi:10.1038/s41746-025-02237-2

2. Perslev M, Darkner S, Kempfner L, Nikolic M, Jennum PJ, Igel C. U-Sleep: resilient high-frequency sleep staging. *Npj Digit Med.* 2021;4(1):72. doi:10.1038/s41746-021-00440-5

3. Phan H, Mikkelsen K, Chen OY, Koch P, Mertins A, De Vos M. SleepTransformer: Automatic Sleep Staging With Interpretability and Uncertainty Quantification. *IEEE Trans Biomed Eng*. 2022;69(8):2456-2467. doi:10.1109/TBME.2022.3147187

4. Salfi F, Corigliano D, Amicucci G, et al. The potential of ensemble-based automated sleep staging on single-channel EEG signal from a wearable device. *bioRxiv*. Preprint posted online July 24, 2025:2025.07.21.665164. doi:10.1101/2025.07.21.665164

5. Lee YJ, Lee JY, Cho JH, Choi JH. Interrater reliability of sleep stage scoring: a meta-analysis. *J Clin Sleep Med JCSM Off Publ Am Acad Sleep Med*. 2022;18(1):193-202. doi:10.5664/jcsm.9538
